# Estimating Counterfactual Placebo HIV Incidence in HIV Prevention Trials Without Placebo Arms Based on Markers of HIV Exposure

**DOI:** 10.1101/2022.05.06.22274780

**Authors:** Yifan Zhu, Fei Gao, David Glidden, Deborah Donnell, Holly Janes

## Abstract

Given recent advances in HIV prevention, future trials of many experimental interventions are likely to be “active-controlled” designs, whereby HIV negative individuals are randomized to the experimental intervention or an active control known to be effective based on a historical trial. The efficacy of the experimental intervention to prevent HIV infection relative to placebo cannot be evaluated directly based on the trial data alone. One approach that has been proposed is to leverage an HIV exposure marker, such as incident rectal gonorrhea which is highly correlated with HIV infection in populations of men who have sex with men (MSM). Assuming we can fit a model associating HIV incidence and incidence of the exposure marker, based on data from multiple historical studies, incidence of the marker in the active-controlled trial population can be used to infer the HIV incidence that would have been observed had a placebo arm been included, i.e. a “counterfactual placebo”, and to evaluate efficacy of the experimental intervention relative to this counterfactual placebo. We formalize this approach and articulate the underlying assumptions, develop an estimation approach and evaluate its performance in finite samples, and discuss the implications of our findings for future development and application of the approach in HIV prevention. Improved HIV exposure markers and careful assessment of assumptions and study of their violation are needed before the approach is applied in practice.

## 1. Introduction

The last decade has seen dramatic success in the field of HIV prevention, with antiretrovirals (ARVs) proven highly effective for HIV prevention when used to treat HIV-infected persons to prevent transmission of infection to partners, and when used by healthy HIV-uninfected persons as pre-exposure prophylaxis (PrEP) (Saag et al., 2020). Multiple PrEP products have been found effective and are in various stages along the regulatory approval path (Chou et al., 2019; Baeten et al., 2016; Nel et al., 2016; Harel et al., 2019; Mayer et al., 2020; Landovitz et al., 2021; Delany, 2021). Despite these successes, HIV remains a major threat to global health. The Joint United Nations Programme on HIV/AIDS (UNAIDS) estimated that 1.5 million people were newly infected with HIV in 2018, with much of the disease burden concentrated in southern Africa (The Joint United Nations Programme on HIV/AIDS, 2021). There remain considerable barriers to implementing existing prevention interventions, involving socio-behavioral, cultural, and limited resource challenges (Sugarman, 2014; Knight et al., 2016). Additional biomedical prevention interventions will be needed to bring a halt to the HIV epidemic.

A variety of new HIV prevention interventions are in various stages of clinical development, including alternative PrEP agents or delivery mechanisms, vaccines, additional on-demand products, and broadly-neutralizing monoclonal antibodies (AIDS Vaccine Advocacy Coalition, 2022). While a traditional placebo-controlled randomized trial that enrolls HIV-uninfected individuals and follows them for incident HIV infection has historically been required for regulatory approval of a new intervention, for new interventions in the same “class” as an intervention already proven effective, e.g. for new PrEP agents, a placebo-controlled design is likely unethical (World Health Organization, 2021). Current and future trials are likely to be “active-controlled” trials, wherein HIV-uninfected participants are randomized to the experimental intervention or an existing “active control” intervention. Even for new interventions in as-yet-unproven classes, e.g. HIV vaccines, there may be future circumstances in which an active-controlled design is necessary.

The fundamental challenge of an active-controlled trial is that absolute prevention efficacy of the experimental intervention, defined as the reduction in HIV incidence for the intervention relative to placebo, cannot be evaluated based on the trial data alone. A traditional approach to deal with this issue is to use data from a historical placebo-controlled trial of the active control to set a “margin” for establishing non-inferiority or superiority of the experimental intervention, based on the assumption that efficacy of the active control established in the historical trial can be carried over to the new trial (James Hung et al., 2003; Fleming, 2008). Such an approach is challenging to employ in HIV prevention, since many interventions are highly user-dependent and efficacy in the historical trial may not apply to the current trial (Grobler and Abdool Karim, 2012; Cutrell et al., 2017; Hanscom et al., 2019). In addition, while efficacy in a non-inferiority design is quantified by *relative efficacy* comparing the experimental and active control arms, absolute efficacy is arguably the parameter of most interest for an experimental intervention that is intended as an addition to the portfolio of HIV prevention options (Glidden, 2019; Glidden et al., 2020). A final challenge with the non-inferiority approach is sample size: non-inferiority trials generally require larger sample sizes than placebo-controlled trials, especially if the active control is highly effective. Developing alternative approaches to evaluating efficacy of a new HIV prevention intervention is therefore a priority.

One approach that has been proposed is to use a marker of HIV exposure as a proxy to infer “counterfactual placebo” HIV incidence, i.e. the incidence that would have been observed had a placebo arm been included in the active-controlled trial. This requires establishing first the association between HIV incidence and incidence of an HIV exposure marker in the absence of intervention, estimated based on historical data. Under the assumption that an intervention in the active-controlled trial does not affect the HIV exposure marker, incidence of the marker in the active-controlled trial can be used to estimate the counterfactual placebo HIV incidence for the trial population. Figure 1 illustrates the concept. The approach has been proposed in concept (Mullick and Murray, 2019; Murray, 2019b) and widely discussed in the HIV prevention field (Murray, 2019a; Janes et al., 2019; Murray, 2019b; Follmann, 2019; Glidden, 2019; Cohen and Donnell, 2019; Glidden, 2020; Glidden et al., 2020; The Forum for Collaborative Research, 2021). The specific proposal has been to use incident rectal gonorrhea infection (RGC) as the marker of HIV exposure for the MSM population. This is based on a body of observational data suggesting that the incidences of these two sexually-transmitted infections are highly correlated; a recent meta-analysis found a coefficient of determination of 0.87 between incidences of HIV and RGC (Mullick and Murray, 2019). US Food and Drug Administration advisory committees reviewing new PrEP agents have appeared to support the approach (US Food and Drug Administration, 2018), and the FDA itself has appeared to endorse the approach in guidance to industry.

**Figure 1:**
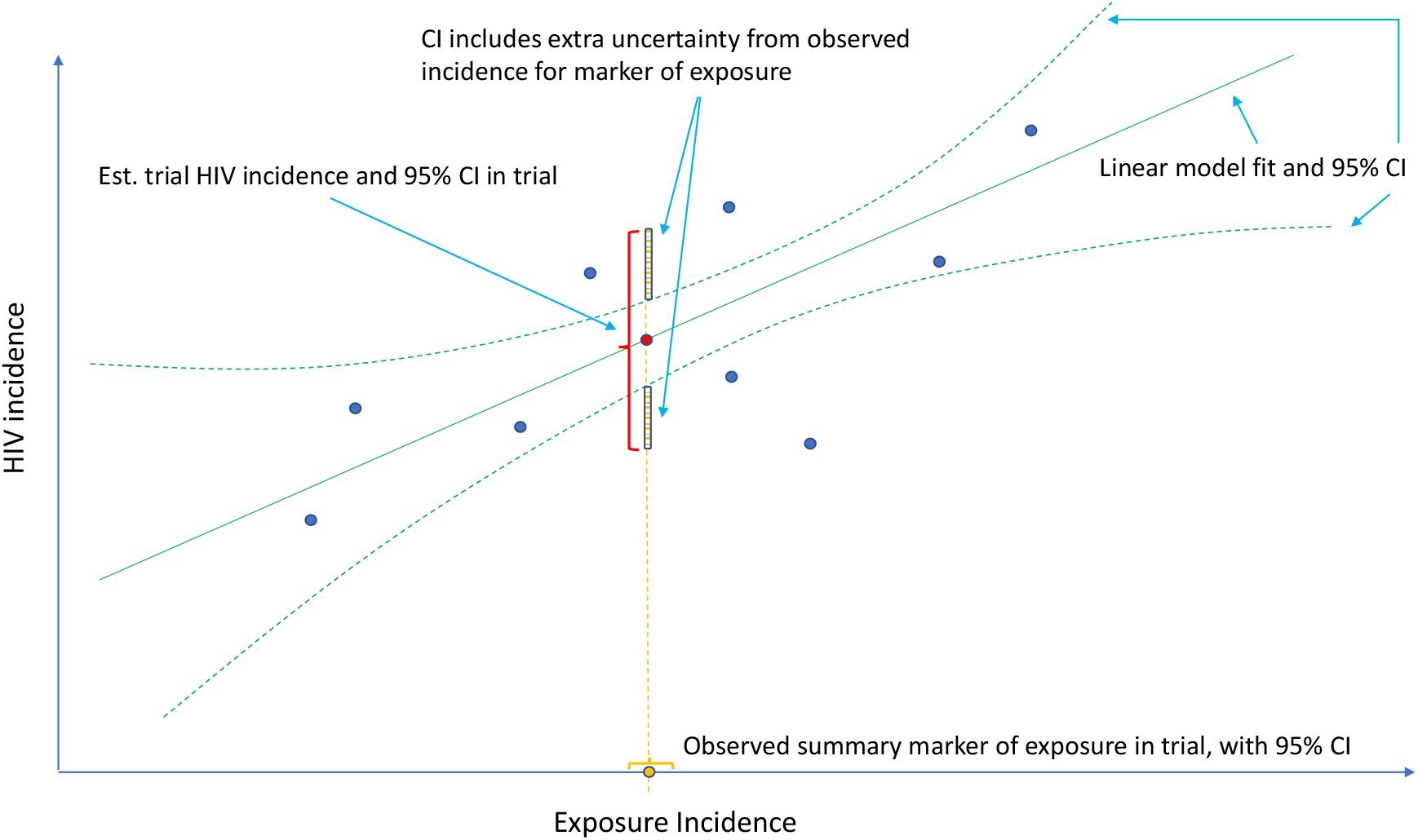
Estimation of counterfactual HIV incidence based on an HIV exposure marker. Green solid and dashed curves correspond to the fitted model associating HIV and exposure marker incidences with an associated pointwise 95% confidence interval, based on a set of external cohorts reporting HIV and exposure marker incidence rates (dark blue dots). Given the exposure marker incidence in the active-controlled trial (yellow dot), counterfactual placebo HIV incidence is estimated with use of the fitted model (red dot). The 95% confidence interval for the counterfactual placebo incidence captures uncertainty due to the model fit and uncertainty in the exposure marker incidence.

In this paper, we articulate a statistical framework for inferring counterfactual placebo HIV incidence for an active-controlled trial using a marker of HIV exposure. We describe an estimation approach and articulate the assumptions under which this approach produces unbiased estimates. We conduct a simulation study designed to mimic closely the specific proposal to use RGC incidence to infer counterfactual placebo HIV incidence, and evaluate the performance of the methods under idealized conditions in which all the underlying assumptions are satisfied. We highlight the limitations of the proposed approach by discussing the validity of the assumptions and challenges with assessing them, and discuss implications for use of the approach in future HIV prevention trials. Our purpose is to guide further discussion around the potential value of RGC as an HIV exposure marker, and to guide the field towards markers that may more readily satisfy the requisite assumptions for reliable inference.

## 2. Methods

### 2.1 Setting and notation

Let *X* indicate the intervention to prevent HIV infection, where *X* = 0 denotes placebo and *X* = 1, 2, …, *K* denote one or more randomly-assigned interventions in the active-controlled trial. Let *Y* denote the HIV infection event time, and 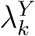 denote the HIV incidence rate for subjects randomized to *X* = *k* in the trial. The quantity of primary interest is the prevention efficacy (PE) of intervention *k*, given by

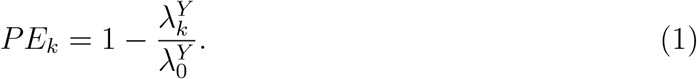

#### Remark 1

We emphasize that efficacy is always evaluated against a backdrop of the current HIV Standard of Prevention for the target population, consisting of proven and available means of HIV prevention (World Health Organization, 2021). Therefore, 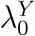, hereafter “placebo incidence”, is defined as the HIV incidence in a counterfactual world where trial participants are randomized to receive a placebo in addition to the Standard of Prevention. Moreover, interpretation of 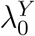 and of prevention efficacy (*PE*_*k*_) is specific to the chosen background Standard of Prevention.

#### Remark 2

PE may be measured on different scales. Generally, we define PE as

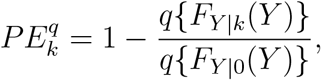

where *F*_*Y* |*k*_(*y*) is the distribution function of the HIV event time *Y* given *X* = *k*, and *q* is a known function. For example, we may define PE based on the instantaneous hazard or cumulative incidence up to a given landmark time. The methods discussed herein generalize directly to estimating the general form of PE.

In a randomized placebo-controlled trial, 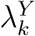 and 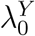 can be estimated based on the observed trial data. However, in a randomized active-controlled trial without a placeboarm, while 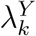 can be estimated based on the observed data, 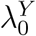, and therefore *PE*_*k*_, cannot be estimated directly. The proposal we formalize is to make use of data from external cohorts to establish a relationship between HIV and an exposure marker, so as to infer 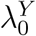.

### 2.2 Assumptions

Let 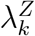 denote the incidence of HIV exposure marker *Z* in the active-controlled trial population, given randomization to intervention *X* = *k*. We wish to establish a relationship between 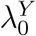 and 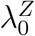 based on data from *M* external cohorts, each of which is conducted under a specific standard of HIV prevention. This standard of prevention likely differs across populations due to local circumstances and due the emergence of new and effective means of HIV prevention over time. We refer to these as “placebo” incidences for simplicity. For external cohort *m*, let 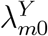 and 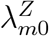 be the incidences of HIV and the exposure marker, respectively. Let 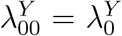 and 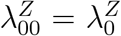 denote the incidence parameters for the active-controlled trial population itself.

We parameterize the relationship between the incidences of HIV and the exposure marker as follows:

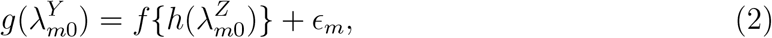

for *m* = 0, …, *M*, where *g*(·) and *h*(·) are known link functions, *g*(·) is an invertible function, *f* (·) is an unknown regression function that can be either parametric or non-parametric, and *ϵ*_*m*_ is an i.i.d. mean-zero error term. Importantly, we view 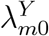 and 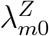 as random variables: there is variability in the placebo incidence rates across cohorts due to different compositions of individuals with heterogeneous HIV risks, and due to differences in the standard of prevention. The link-functions *g*(·) and *h*(·) should be those appropriate for non-negative incidence functions, e.g. logit or log links.

We make the following assumptions.

#### Assumption 1

Regression model (2) describes a general relationship between placebo HIV and exposure marker incidence rates that holds across external cohorts and for the active-controlled trial population.

#### Assumption 2

An unbiased estimate of *f* can be obtained based on observed incidences 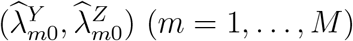 (and their reported variances) in the external cohorts. The estimate is denoted by 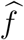.

#### Assumption 3

The exposure marker incidence is not modified by randomization to active intervention *X* = *k*, i.e., 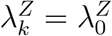.

Assumption 1 requires a certain level of similarity across the trial and all external cohorts: while the incidences of HIV and exposure markers may differ, the *association* between the incidence rates is assumed to be constant. In evaluating Assumption 1, one must consider carefully the background Standard of Prevention for the active-controlled trial population, and whether any element of this prevention package may influence the relationship between HIV and the exposure marker. For example, oral PrEP is known to reduce HIV but does not have a biological effect on RGC or other non-HIV STIs (Mayer et al., 2020), even though it may have an effect in terms of behavioral “risk disinhibition” (Traeger et al., 2019). Therefore, if the trial Standard of Prevention does not include oral PrEP, the external cohorts should be drawn from populations without access to PrEP– which suggests that they will be older cohort studies, before the advent of PrEP. Elements of the Standard of Prevention such as condoms and risk reduction counseling may be reasonably expected to influence incidences of HIV and RGC, but not to modify their association, and therefore may not be critical to consider in evaluating external cohorts. Other potential effect modifiers of the association between HIV and the exposure marker need to be considered, including subject demographics, behaviors, and features of the local HIV epidemic such as population prevalence of HIV. Another consideration is whether blinding may influence the relationship between HIV and the exposure marker. While the counterfactual placebo arm is (conceptually) blinded, the external cohorts may not be.

Assumption 2 indicates that the relationship between HIV and exposure marker incidences can be consistently estimated by observed incidence rates from the external cohorts. This assumption may not be trivial since the reported incidences 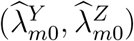 are associated with additional variability so (2) may not generally hold with 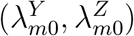 replaced by 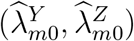 This assumption will be discussed below as it pertains to our estimation approach.

Assumption 3 stipulates that the HIV exposure incidence for arm *X* = *k* of the trial is the same as that under placebo. In evaluating this assumption one must consider whether the intervention, or elements of the trial Standard of Prevention, may modify the incidence of the exposure marker. Whether or not the active-controlled trial is blinded is also relevant, since knowledge of receipt of intervention may modify behavior. Note that Assumption 3 need only hold for one intervention in the trial.

Consider the testability of these assumptions. Assumption 3 can be partially evaluated, ideally in the context of an historical randomized, placebo-controlled trial of the intervention with the exposure marker collected as an endpoint, and less reliably based on an historical prospective cohort study. In either case, however, the historical data inform on the validity of Assumption 3 only as long as the effect observed in the historical study is assumed to apply to the trial population, where placebo incidence of *Z* is not observed. Assumption 2 is specific to the estimation approach; given a form for model (2), the assumption may or may not hold depending on whether the estimation approach would adequately account for the additional variability of 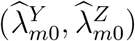 given the true incidences 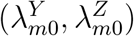 In the following sections, we will discuss validity and violation of Assumption 2 in a special case of model (2). Assumption 1 is not fully testable as it pertains to the unobservable counterfactual placebo incidence in the trial population; the correct specification of model (2) for the external cohorts may be evaluated but it cannot be evaluated for the trial population.

Under Assumptions 1-3, we may estimate counterfactual placebo HIV incidence by

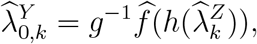

where 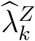 is the observed exposure marker incidence in the trial arm randomized to intervention *k*. Importantly, the uncertainty of the estimated counterfactual placebo HIV incidence is comprised of the uncertainty due to fitting the regression model, 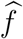, and the uncertainty in the exposure marker incidence, 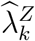, as illustrated in Figure 1. Prevention efficacy, *PE*_*k*_, can then be estimated by

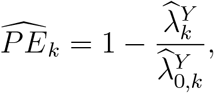

where 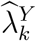 is the estimated HIV incidence among those randomized to intervention *k* in the active-controlled trial.

#### Remark 3

If we assume that the exposure marker incidence is not modified by a collection of interventions *X* ∈ 𝒦 in the active-controlled trial, we may use the observed exposure marker incidence among trial participants who received intervention 𝒦 to obtain an estimated counterfactual placebo HIV incidence with increased precision.

### 2.3 Bivariate linkage model

In this and the following sections, we focus on a special case of model (2) for which we discuss validity and violation of Assumption 2. Specifically, we assume a bivariate normal distribution for logit-transformed HIV and exposure marker incidence rates. Write 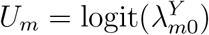 and 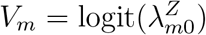. We assume

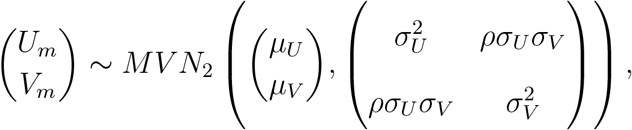

for *m* = 1, …, *M*, where *ρ* ∈ (0, 1) measures the magnitude of association between *U*_*m*_ and *V*_*m*_. Under this model, model (2) is specified by

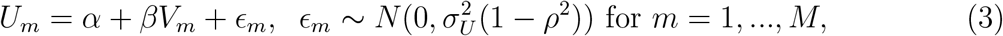

where *α* = *μ*_*U*_ *− ρμ*_*V*_ *σ*_*U*_ */σ*_*V*_ and *β* = *ρσ*_*U*_ */σ*_*V*_.

We assume that the estimated incidence rates from the external cohorts 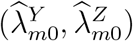 are independent given the true incidence rates 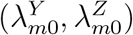. Such a conditional independence assumption has been commonly adopted in related bivariate outcome meta-analysis literature (Van Houwelingen et al., 2002; Reitsma et al., 2005). We will develop methods based on this conditional independence assumption, and delay discussion on violation of such assumption to Section 2.6. We write 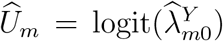 and 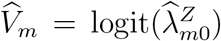. The resultant conditional joint distribution is given by

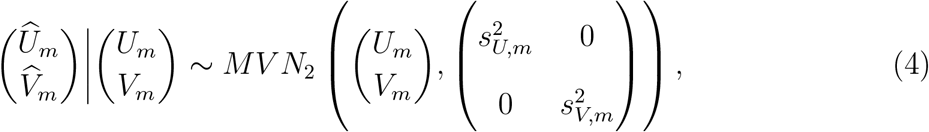

where 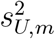 and 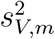 are the conditional variances of 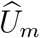 and 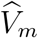 given (*U*_*m*_, *V*_*m*_), respecttively.

Combining (3) and (4), the distribution for 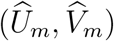 is given by

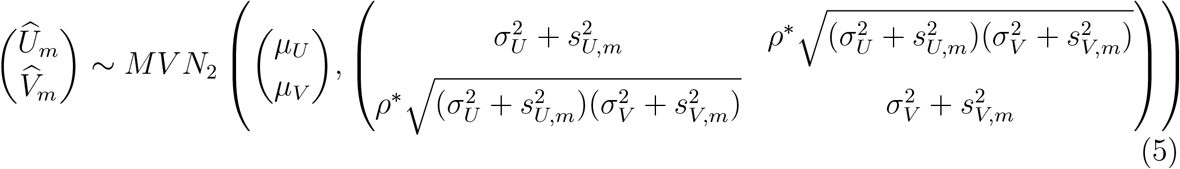

where 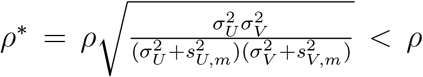 That is, the association between 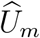 and 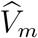 is weaker than that between *U*_*m*_ and *V*_*m*_; it is attenuated given the additional variability of 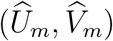 given (*U*_*m*_, *V*_*m*_). It follows that the conditional distribution of 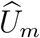 given 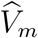 is given by

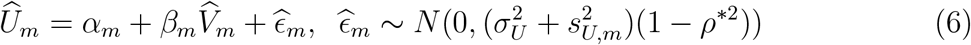

where 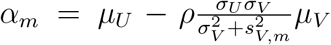 and 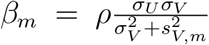 are cohort-specific. This suggests that naively linearly regressing 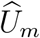 on 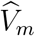 may not produce an unbiased estimate of the regression function, *f*.

### 2.4 Maximum Likelihood Estimation

Under the bivariate linkage model, expression (5) specifies the likelihood of the observed data 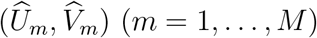 given parameters 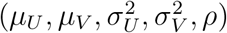. An unbiased estimator for 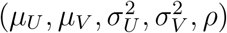 may be obtained through maximizing the likelihood function, so as to produce an unbiased estimator for *f* that satisfies Assumption 2. Specifically, the log-likelihood function is given by

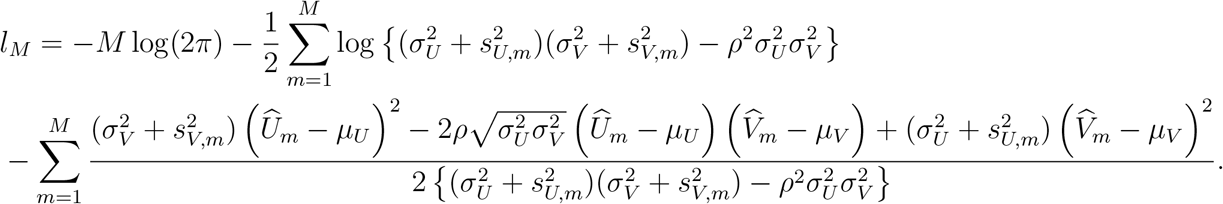

In practice, we may replace 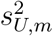and 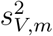 in the log-likelihood function by their estimates from the external cohorts. Direct maximization of the log-likelihood function may not be stable, and therefore we develop an EM algorithm for computation, with details in Supplementary Materials. Let 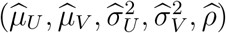 denote the resultant maximum likelihood estimates.

#### 2.4.1 Counterfactual Placebo HIV Incidence Estimation

Based on the estimated parameters, estimate the counterfactual placebo HIV incidence in the trial, 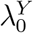, based on the observed exposure marker incidence 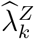 Write 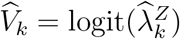 The conditional mean of *U*_0_ *≡* logit 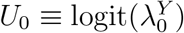 given 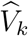 is given by

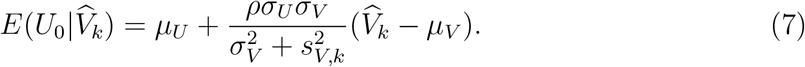

That is, we estimate *U*_0_ using an estimator that replaces the parameters in (7) by their estimates, given by

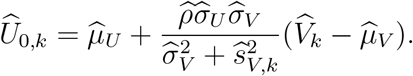

The variance of this estimator is computed by applying the delta method based on the covariance matrix of 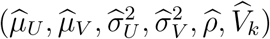 noting that 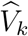 and 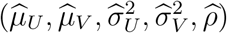 are independent. The formula is given in the supplementary material.

#### 2.4.2 Estimating Prevention Efficacy

A natural estimate for PE as defined in (1) is obtained by replacing 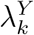 and 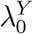 by their respective estimators. Specifically, we estimate PE by

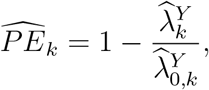

where 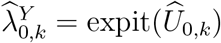. The asymptomatic variance of 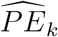 is estimated using the delta-method, given that 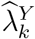 and 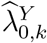 are independent and asymptotically normally distributed given *V*_*k*_ Details are given in the supplementary materials.

### 2.5 Working regression model

While maximum likelihood estimation yields consistent and efficient parameter estimates under correct model specification, it may not be stable when the number of external cohorts is small, e.g., *M <* 20, as suggested by our simulations. Therefore, we consider alternatively fitting a working regression model

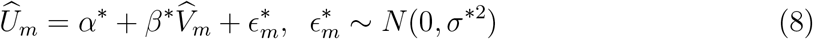

that is generally mis-specified relative to the true model (6) since a common intercept and slope are assumed across external cohorts. Indeed, (8) is correctly specified if and only 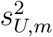 and 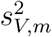 are the same for all *m* = 1, …, *M*. We denote the fitted working model parameters by 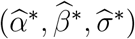. The estimated regression function, 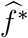, based on 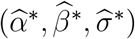, may approximate *f* well enough to provide adequate inference about counterfactual placebo HIV incidence. We evaluate in simulations the bias associated with fitting the mis-specified working model.

Given working model estimates 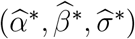, counterfactual placebo HIV incidence is estimated by

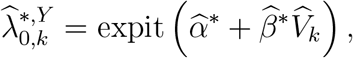

and PE is estimated by

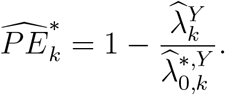

The confidence interval for 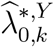 is constructed based on a t-distribution approximation, given an asymptotic variance derived using the delta method. The analytical variance of 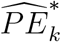 is non-trivial, as it involves the ratio between asymptotically normally-distributed 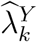 and approximately t-distributed 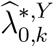, see e.g., Nadarajah and Dey (2006); Nadarajah (2006). Therefore, to quantify uncertainty in 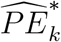 we apply the bootstrap whereby *M* external cohorts are sampled with replacement.

### 2.6 Violation of the conditional independence assumption

Previous sections assumed 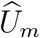 and 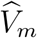 are conditionally independent given (*U*_*m*_, *V*_*m*_) with the conditional distribution given in (4). In practice, such conditional independence may not hold and a more general conditional distribution is

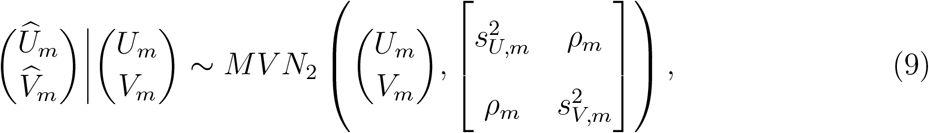

where *ρ*_*m*_ measures the cohort-specific association between 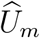and 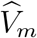, based on the fact that the two outcomes are measured on the same participants. The joint distribution of 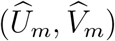 then takes the form

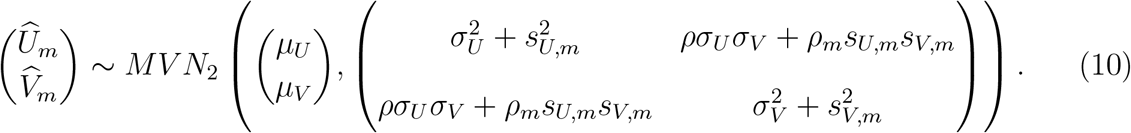

One challenge with applying this more general distribution in practice is that typically *ρ*_*m*_ is not reported by external cohort studies. Either sensitivity analyses that incorporate empirical knowledge of the degree of conditional dependence or bootstrapping of individual-level data (Daniels and Hughes, 1997) is recommended to estimate the within-study correlation (Riley, 2009) and to perform inference with model (10). However, even with *ρ*_*m*_ estimates in hand, there are technical challenges associated with performing inference based on model (10) (see Daniels and Hughes (1997); Riley (2009); Riley et al. (2015); Hong et al. (2018)). Therefore, we leave estimation under the more general conditional dependence model for future research, and evaluate estimation using model (5) that assumes conditional independence. In simulations described below, we evaluate the performance of this estimation when the conditional independence assumption is violated.

## 3. Simulation Studies

### 3.1 Simulation methods

We designed simulation studies to reflect the HIV prevention context, and the specific proposal to use RGC as a marker of HIV exposure. The model parameters in (5) were estimated based on published studies reporting both HIV and RGC incidence for MSM populations, summarized in Supplementary Table 1. The MLE estimates are 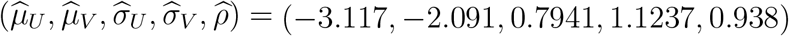 and serve as true model parameters in the simulations. The estimated correlation parameter *ρ* is high. To evaluate the influence of the correlation parameter on the approach, we also consider a moderate correlation scenario with *ρ* = 0.5. The number of external cohorts *M* is set at 10 or 20, reflecting the practical reality that generally only a small number of external studies will be available with paired HIV and exposure marker data. We also explore large *M* scenarios (*M* = 50, 100, 1000) to verify expected large-sample operating characteristics. External cohort person-time is uniformly distributed between 200 and 5000 person-years. We assume a single arm in the trial (*K* = 1) for conciseness, with a total follow-up time *n*_*x*_ = 500, 1000, 2000, or 4000 person-years. PE is assumed to be 30%, 60% or 75%. Placebo HIV incidence is assumed to be 3, 4.5, or 6 per 100 person-years. Both the likelihood-based and working regression model estimation approaches are evaluated. Results are summarized across 1000 simulations.

### 3.2 Simulation results

Table 1 summarizes the performance of estimated counterfactual placebo HIV incidence across simulation scenarios. We show results with *M* = 10 and 20 for the working regression model estimation approach, and we show results with *M* = 20 and 50 for the likelihood-based estimation approach, as likelihood-based estimation requires sufficiently large *M* to ensure numerical stability given that it requires estimating more parameters. For both estimation approaches, we find that high correlation between HIV and the exposure marker (*ρ* = 0.938) yields accurate and precise estimation, as evidenced by low bias and confidence intervals with reasonable width and close to nominal coverage. For example, with working-model-based estimation, *M* = 10 external cohorts, and 2,000 person-years trial follow-up, true counterfactual HIV incidence of 4.5% is estimated with less than 1% bias, and the 95% confidence interval has 96% coverage and is narrow, 3.6% to 5.5%. Even with modest correlation (*ρ* = 0.5), low bias and nominal coverage rates are seen, given a sufficient number of external cohorts (*M* ⩾ 20). Performance is only minimally impacted by the size of the active arm of the trial. Performance of the working regression estimation approach is comparable to that of likelihood-based estimation in the settings with *M* = 20, while it performs worse for large *M*, with confidence intervals that are overly conservative (results not shown).

**Table 1:**
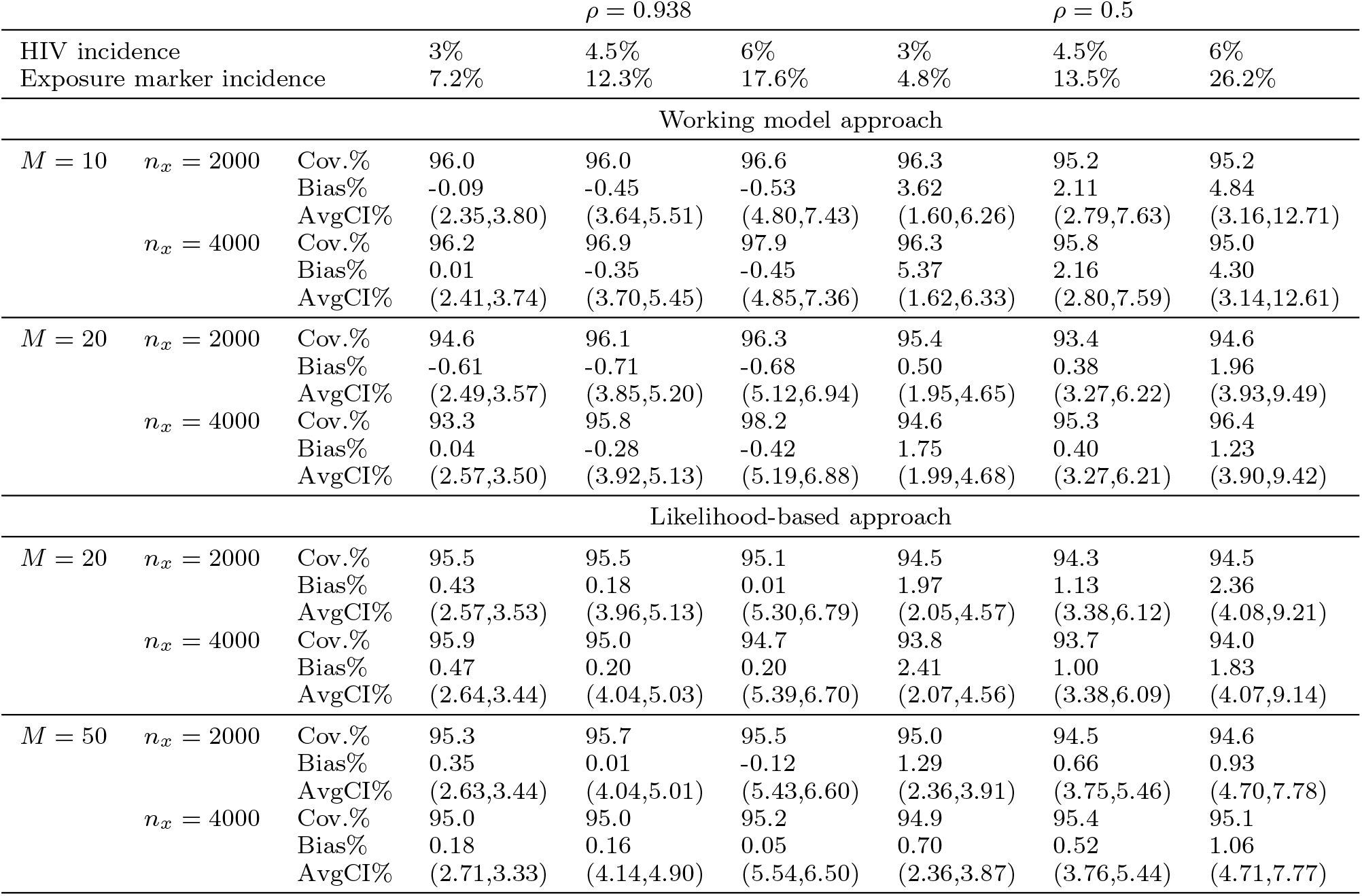
Percent bias, empirical coverage, and average confidence intervals (CIs) for estimated counterfactual placebo HIV incidence, based on *M* external cohorts used to estimate the association between HIV and an exposure biomarker with correlation *ρ*. A total of *n*_*x*_ person-years follow-up accrue in the active arm of the trial. Counterfactual placebo HIV incidence varies. Performance is shown for working model and likelihood-based estimation approaches.

The performance of estimates of PE based on an active-controlled trial with *n*_*x*_ = 2000 is shown in Table 2. When *ρ* is large, PE can be estimated with low bias with confidence intervals with near-nominal coverage and reasonable width, even with *M* = 10 external cohorts. For example, when PE is 60% against a 4.5% placebo HIV incidence, with 10 external cohorts there is less than 1% bias and the nominal 95% confidence interval has 94.7% coverage and on average extends from 42.3% to 72.9%. However, with modest *ρ*, PE may be estimated with larger bias and confidence intervals slightly under-cover with small *M* (*M* = 10). For the same *M*, modest *ρ* generally yields a wider confidence interval. For scenarios with high *ρ*, PE can be estimated with better precision when the placebo HIV incidence rate is higher, resulting from the fact that incidences can be more precisely estimated when there are more incident events. With modest *ρ*, however, the confidence intervals are wider when the placebo HIV incidence is 3% or 6% compared to 4.5%. This is because, with moderate *ρ*, the variability of the PE estimate is more dominated by the variability of the estimated counterfactual placebo HIV incidence, which is larger when the placebo HIV incidence is further from the mean HIV incidence across the external cohorts, as illustrated in Figure 1. With *M* = 20, the average confidence interval widths are similar for the working regression approach and the likelihood-based approach, although the confidence intervals from the likelihood-based approach have slightly lower coverage in some cases and are shifted upwards. Performance based on a smaller active-controlled trial with *n*_*x*_ = 1000 person-years follow-up is shown in Supplementary Table 2; confidence intervals are wider but coverage rates and bias are only minimally worse.

**Table 2:** Percent bias, empirical coverage, and average 95% confidence intervals (CIs) for estimates of prevention efficacy (PE) based on *M* external cohorts used to estimate the association between HIV and an exposure biomarker with correlation *ρ*. A total of *n*_*x*_ = 2000 person-years follow-up accrue in the active arm of the trial. Counterfactual placebo HIV incidence and true PE vary. Performance is shown for working model and likelihood-based estimation approaches.

| $M$ | | PE estimate | $\rho = 0.938$ | | | $\rho = 0.5$ | | |
| --- | --- | --- | --- | --- | --- | --- | --- | --- |
| HIV incidence |  |  | 3% | 4.5% | 6% | 3% | 4.5% | 6% |
| Exposure marker incidence |  |  | 7.2% | 12.3% | 17.6% | 4.8% | 13.5% | 26.2% |
| Working model approach |  |  |  |  |  |  |  |  |
| $PE = 30\%$<br>$n_x = 2000$ | 10 | Bias% | -0.06 | -1.04 | -1.30 | 8.29 | 4.26 | 9.79 |
|  |  | Cov.% | 95.2 | 93.6 | 94.0 | 91.9 | 93.0 | 91.4 |
|  |  | AvgCI% | (-1.5,52.7) | (4.6,48.2) | (6.7,47.2) | (-63.0,62.7) | (-21.4,56.3) | (-63.2,60.4) |
|  | 20 | Bias% | -1.41 | -1.69 | -1.67 | 2.09 | 0.49 | 3.27 |
|  |  | Cov.% | 95.6 | 94.3 | 94.0 | 94.2 | 92.8 | 93.8 |
|  |  | AvgCI% | (1.4,51.3) | (7.5,47.4) | (10.5,45.9) | (-21.7,56.8) | (-3.8,52.3) | (-15.6,54.8) |
| $PE = 60\%$<br>$n_x = 2000$ | 10 | Bias% | -0.06 | -0.30 | -0.36 | 2.33 | 1.38 | 3.08 |
|  |  | Cov.% | 94.8 | 94.7 | 94.2 | 92.3 | 93.8 | 92.0 |
|  |  | AvgCI% | (37.8,75.7) | (42.3,72.9) | (43.7,71.7) | (4.5,80.4) | (28.5,76.7) | (4.3,78.2) |
|  | 20 | Bias% | -0.41 | -0.47 | -0.46 | 0.33 | 0.25 | 1.28 |
|  |  | Cov.% | 95.7 | 94.9 | 95.4 | 94.8 | 94.7 | 94.8 |
|  |  | AvgCI% | (39.6,75.3) | (44.1,72.7) | (46.2,71.4) | (27.0,77.7) | (38.0,74.6) | (33.1,75.7) |
| $PE = 75\%$<br>$n_x = 2000$ | 10 | Bias% | -0.03 | -0.16 | -0.19 | 1.18 | 0.62 | 1.42 |
|  |  | Cov.% | 94.6 | 94.5 | 94.8 | 93.8 | 92.8 | 92.7 |
|  |  | AvgCI% | (58.1,86.8) | (61.6,84.7) | (62.9,83.8) | (37.8,89.0) | (53.4,86.6) | (39.5,87.1) |
|  | 20 | Bias% | -0.22 | -0.25 | -0.24 | 0.22 | 0.15 | 0.64 |
|  |  | Cov.% | 95.1 | 94.6 | 94.5 | 95.2 | 94.5 | 94.4 |
|  |  | AvgCI% | (59.7,86.8) | (62.6,84.7) | (64.7,83.8) | (52.4,87.9) | (59.2,85.6) | (56.8,85.9) |
| Likelihood-based approach |  |  |  |  |  |  |  |  |
| $PE = 30\%$<br>$n_x = 2000$ | 20 | Bias% | -0.65 | -0.70 | -0.87 | -4.86 | -3.13 | -5.11 |
|  |  | Cov.% | 93.8 | 94.9 | 94.5 | 92.8 | 93.7 | 92.8 |
|  |  | AvgCI% | (6.3,53.3) | (10.7,48.9) | (12.9,46.6) | (-6.3,63.4) | (2.6,55.5) | (-3.3,60.2) |
|  | 50 | Bias% | -0.75 | -1.07 | -0.43 | -0.73 | -0.45 | -2.23 |
|  |  | Cov.% | 94.1 | 94.7 | 94.8 | 93.8 | 94.0 | 94.3 |
|  |  | AvgCI% | (6.9,52.7) | (11.1,48.3) | (13.7,46.0) | (2.3,57.2) | (8.4,51.3) | (6.3,52.4) |
| $PE = 60\%$<br>$n_x = 2000$ | 20 | Bias% | -0.32 | -0.01 | -0.36 | -1.54 | -0.91 | -1.35 |
|  |  | Cov.% | 93.4 | 94.8 | 94.8 | 92.5 | 93.5 | 93.4 |
|  |  | AvgCI% | (42.8,76.8) | (46.1,73.8) | (47.6,72.0) | (36.3,81.8) | (41.9,77.0) | (39.3,79.1) |
|  | 50 | Bias% | -0.37 | -0.35 | 0.00 | -0.22 | -0.21 | -0.79 |
|  |  | Cov.% | 94.5 | 94.1 | 94.6 | 93.6 | 94.2 | 94.6 |
|  |  | AvgCI% | (43.0,76.5) | (46.1,73.4) | (48.2,71.9) | (41.0,78.8) | (44.9,74.9) | (44.2,74.8) |
| $PE = 75\%$<br>$n_x = 2000$ | 20 | Bias% | 0.03 | 0.03 | -0.12 | -0.65 | -0.41 | -0.65 |
|  |  | Cov.% | 93.2 | 94.0 | 94.4 | 92.0 | 93.2 | 93.2 |
|  |  | AvgCI% | (61.9,88.1) | (64.3,85.7) | (65.5,84.3) | (58.3,90.8) | (62.0,87.4) | (60.8,88.2) |
|  | 50 | Bias% | -0.11 | -0.19 | -0.01 | -0.21 | -0.17 | -0.44 |
|  |  | Cov.% | 93.5 | 93.6 | 94.6 | 93.5 | 94.0 | 94.2 |
|  |  | AvgCI% | (61.9,87.9) | (64.2,85.5) | (64.8,84.2) | (60.7,89.0) | (63.5,86.2) | (63.6,85.8) |

Figure 2 shows power for testing *H*_0_ : *PE* = 30% under the design alternative *H*_*a*_ : *PE* = 60% and *n*_*x*_ = 2000, as a function of *M*. For reference, the figure also shows the size of a placebo arm that would be required to achieve the same power using a traditional randomized placebo-controlled trial. For example, with moderate *ρ* and 6% placebo HIV incidence, with an active arm with 2,000 person years and 50 external cohorts, power to detect 60% PE is just above 90%, roughly equivalent to a placebo-controlled trial with 1,817 placebo participant person-years follow-up. For a given combination of *ρ* and 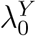, we see that power increases as a function of *M*. However, the rate of increase is higher with moderate vs. high *ρ*: with high *ρ*, the linkage model can already be estimated precisely with small *M*, while with moderate *ρ*, more external cohorts are needed for precise estimation. With an unrealistically large number of external cohorts (*M* = 1000), power is similar for high and moderate *ρ*. For *M* = 10 and 20, we show power for both the working-model-based and likelihood-based estimation approaches, and we observe higher power as expected with likelihood-based estimation. Surprisingly, we find that power from the counterfactual approach can actually exceed that obtained from a placebo-controlled trial with the same active-arm size. The reason is that in our scenarios, incidence of the exposure marker is higher than that of HIV, and thus with a highly correlated exposure marker HIV incidence can be estimated more precisely by leveraging information in the higher-incidence exposure marker. Supplementary Figures 1-3 show analogous results with different active-arm sizes (*n*_*x*_ = 500, 1000 person-years) and a different design alternative, *H*_*a*_ : *PE* = 75%.

**Figure 2:**
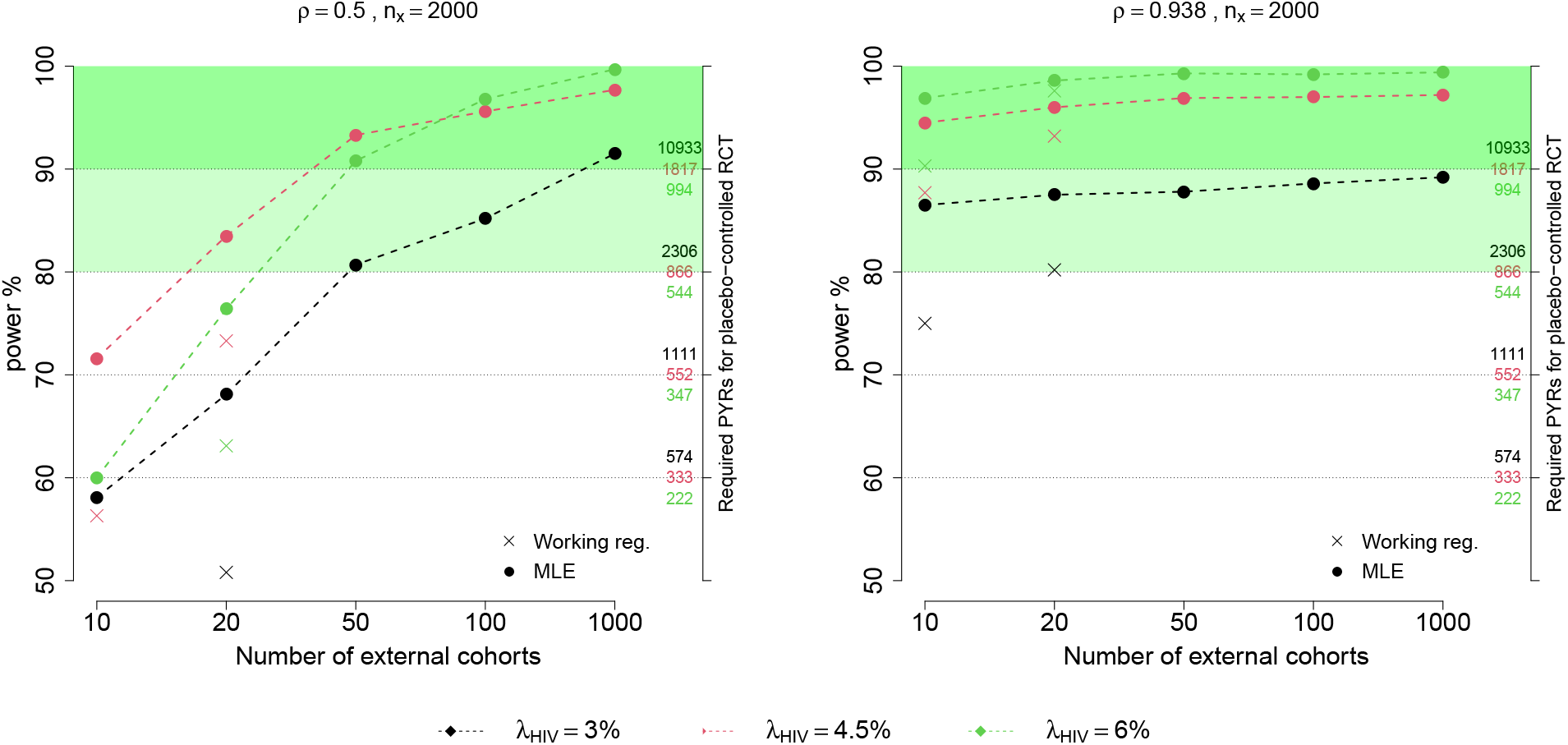
Power for testing *H*_0_ : *PE* = 30% vs. *H*_*a*_ : *PE* = 60% using the counterfactual approach as a function of *M*, the number of external cohorts. Given a fixed active arm size of 2000 person-years, the power based on estimating counterfactual placebo HIV incidence using MLE estimation, with a moderately correlated exposure marker (*ρ* = 0.5; left) or a highly correlated marker (*ρ* = 0.938; right). The size of a placebo arm, in person-years, required to obtain the corresponding power is shown on the right-hand y-axis. The power calculated from the working regression estimation approach is also shown for low *M*.

### 3.3 Impact of violation of the conditional independence assumption

We performed additional simulations in which the conditional independence assumption is violated: the external cohort estimates were generated from (10), where the within-cohort correlations, *ρ*_*m*_, were randomly generated from *U* (0.4, 0.5). As shown in Supplementary Tables 3 and 4, the performance of the counterfactual placebo incidence and PE estimates is similar to what is reported above under conditional independence, suggesting that the estimation is reasonably robust to violation of the conditional independence assumption. Similar robustness has been reported in related bivariate meta-analysis work Riley (2009). One of the conditions that minimizes the impact of conditional dependence is small within-study variation relative to between-study variation, and this condition is largely satisfied under our simulation scenarios.

### 3.4 External cohorts: more cohorts of smaller sizes or fewer cohorts of larger sizes?

In settings where external cohort data are available at a more granular level than at the study level, e.g., when site-level data are available for multi-center studies, a question is whether performance of the counterfactual placebo estimate is improved when the external cohort data are analyzed at the sub-cohort level. We conducted additional simulations to explore this question. In these simulations, each of *M* = 20 cohorts has *L* = 5 sites and site-level HIV and HIV exposure marker incidences follow model (5). The number of person-years for each site is uniformly distributed as *U* (200*/L*, 5000*/L*) = *U* (40, 1000). We compare results based on cohort-level vs. site-level estimation of counterfactual placebo HIV incidence, where the total sample size of the external cohort data is approximately constant. We find that analyzing data at the site level provides more precise inference, as reflected by narrower confidence intervals with similar coverage rates (see Supplementary Table 5).

## 4. Discussion

Advancing HIV prevention, and ultimately stemming the HIV pandemic, will require the advent, evaluation, and dissemination of additional biomedical interventions that prevent HIV acquisition. While active-controlled trials are likely to be the norm for future trials evaluating a wide variety of candidate interventions, they are subject to a fundamental limitation: absolute efficacy of the experimental intervention cannot be evaluated based on the trial data alone. If a marker of HIV exposure can be measured in the trial, and external data are leveraged to model the association between HIV and the exposure marker, we have demonstrated that HIV incidence in a counterfactual placebo arm, and prevention efficacy of the experimental intervention relative to the counterfactual placebo, can be estimated reliably and precisely.

In this paper, we considered idealized conditions where Assumptions 1 and 3 are satisfied and evaluated the performance of statistical approaches where Assumption 2 may or may not hold. We assumed that at least one intervention in the trial does not impact the marker, and that the correct model associating the marker and HIV holds for both the trial population and the external cohorts. These are strong and partially untestable assumptions that deserve careful attention in practice. In particular, correct specification of the model linking HIV with the exposure marker is extremely challenging and there are numerous ways in which this model might be misspecified, including due to omission of covariates that modify the association, incorrect model form, and measurement error in variables. Standard statistical methods may be applied to check for specific types of model misspecification based on data collected from the external cohorts. However, with few external cohorts, power to detect model mis-specification is low. Given some of the assumptions are not fully testable, further research is needed into methods for incorporating uncertainty due to violation of these assumptions. Application of the approach is not warranted until such research is conducted.

In simulation studies, we found that precision of prevention efficacy is impacted by the distribution of HIV incidence across the external cohorts. Specifically, a higher counter-factual placebo HIV incidence may lead to a less precise efficacy estimate if it is further away from the mean HIV incidence across the external cohorts. Therefore, it is desirable to include external cohorts that are expected to have a similar HIV incidence to the active-controlled trial population. A related practical question is whether to include data from an external cohort if its HIV incidence is far away from the expected placebo incidence in the active-controlled trial. Additional research is needed to explore the impact on statistical performance and to guide future trial design and application.

We explored rectal gonorrhea as a marker of HIV exposure for the MSM population. This is the marker that has received the most attention in the HIV prevention community. However, RGC has major limitations as an exposure marker. Recent work has demonstrated that its association with HIV can differ across studies even within the MSM population (Donnell et al., 2021), and has highlighted the numerous ways in which the HIV-RGC association might be difficult to model accurately across cohorts (Glidden, 2019; Cohen and Donnell, 2019; Glidden, 2020; The Forum for Collaborative Research, 2021). It appears likely that markers more fundamentally linked to HIV exposure will be needed to realize the potential of the counterfactual placebo approach.

We developed inferential methods for the simplest setting where only study-level data are available from external cohorts to model the association between HIV and the exposure marker. However, in some settings individual-level data may be available from external cohorts. Our comparison of inference based on study-level vs. site-level data suggests that accuracy and precision may be further improved with individual-level data, and expansion in this direction is warranted.

Relatedly, with only study-level data from external cohorts, the correlation between reported HIV and the exposure marker incidences in the external cohorts is expected to be available only rarely. Accordingly, our estimation approach assumes conditional dependence of HIV and the exposure marker, and our simulation study suggests a degree of robustness to violation of this assumption. We found that the impact of conditional dependence is negligible, mainly because that in our settings between-study variation dominates with-in study variation. Similar results have been suggested in Riley (2009) for bivariate meta-analysis. However, in general, as discussed in Riley (2009), ignoring within-study correlation is expected to yield estimates with inferior statistical properties. Given individual-level data from external cohorts, estimation of the conditional dependence parameter would be feasible and performance is expected to improve.

We considered the statistical literature at large when evaluating whether existing statistical frameworks provided a good fit for our problem. The HIV exposure biomarker has been, at times, confused with a surrogate outcome, which is in fact quite different because a surrogate outcome is one that is impacted by the intervention (Fleming and Powers, 2012), whereas the concept here is that the exposure biomarker is not. It is similar to an “outcome-inducing confounding proxy” variable as defined in the proximal causal learning framework (Tchetgen et al., 2020). However, utilizing this framework would require postulating the existence of an additional “treatment-inducing confounding” proxy variable which is not readily available in the HIV context. In the epidemiological literature, the concept of a “negative control” outcome appears similar in that it is one not impacted by the exposure of interest– but it is chosen to control the potential biases that are shared with the outcome under study (Arnold and Ercumen, 2016). We found that these statistical frameworks did not fit our application and were therefore led to define one suited for purpose. The framework we have developed could have application in other clinical contexts where there exists a proxy outcome associated with the clinical outcome under the control condition, but not impacted by the intervention; and a body of data for estimating the association between proxy outcome and clinical outcome under the control condition.

## Data Availability

All data and computer codes used to generate these in the present study are available upon reasonable request to the authors.

## Acknowledgements

This work was supported by the National Institutes of Health/National Institute of Allergy and Infectious Diseases (NIH/NIAID) through grants R56AI143418 and UM1AI068635 to HJ.

## Supplementary Materials

Supplementary Materials are available with this paper at the Biometrics website on Wiley Online Library.

