## Supplemental materials for "Estimating Counterfactual Placebo HIV Incidence in HIV Prevention Trials Without Placebo Arms Based on Markers of HIV Exposure"

### Supplementary materials

*Details on the EM algorithm*

To maximize the log-likelihood function, we consider an EM algorithm treating $(U_m, V_m)$ as missing data. Specifically, in the M-step, the log-likelihood for $(U_m, V_m)$ $(m = 1, \ldots, M)$ is given by

$$\begin{aligned} l_M =& - M\log(2\pi\sigma_U\sigma_V\sqrt{1-\rho^2}) \\ & - \frac{1}{2(1-\rho^2)}\sum_{m=1}^{M}\left\{\left(\frac{U_m-\mu_U}{\sigma_U}\right)^2 - 2\rho\left(\frac{U_m-\mu_U}{\sigma_U}\right)\left(\frac{V_m-\mu_V}{\sigma_V}\right) + \left(\frac{V_m-\mu_V}{\sigma_V}\right)^2\right\}. \end{aligned}$$

At the $j$th iteration, the maximizer of $(\mu_U, \mu_V, \sigma_U^2, \sigma_V^2, \rho)$ given $(U_m, V_m)$ is given by

$$\begin{aligned} \widehat{\mu}_U^{(j)} =& M^{-1}\sum_{m=1}^{M} U_m, \qquad \widehat{\mu}_V^{(j)} = M^{-1}\sum_{m=1}^{M} V_m, \\ \widehat{\sigma}_U^{2,(j)} =& M^{-1}\sum_{m=1}^{M}(U_m - \widehat{\mu}_U^{(j)})^2, \qquad \widehat{\sigma}_V^{2,(j)} = M^{-1}\sum_{m=1}^{M}(V_m - \widehat{\mu}_V^{(j)})^2, \\ \widehat{\rho}^{(j)} =& \frac{M^{-1}\sum_{m=1}^{M}(U_m - \widehat{\mu}_U^{(j)})(V_m - \widehat{\mu}_V^{(j)})}{\widehat{\sigma}_U^{(j)}\widehat{\sigma}_V^{(j)}}. \end{aligned}$$

In the E-step, we evaluate the conditional expectation of the terms $(U_m, V_m, U_m^2, V_m^2, U_mV_m)$ given the observed data $(\widehat{U}_m, \widehat{V}_m)$ and current parameter values $(\widehat{\mu}_U^{(j)}, \mu_V^{(j)}, \sigma_U^{2,(j)}, \sigma_V^{2,(j)}, \rho^{(j)})$. Note that the joint distribution of $(\widehat{U}_m, \widehat{V}_m, U_m, V_m)^{\mathrm{T}}$ is given by a multivariate normal, with mean $(\mu_U, \mu_V, \mu_U, \mu_V)^{\mathrm{T}}$ and the covariance matrix is given by

$$\begin{pmatrix} \sigma_U^2 & \rho\sigma_U\sigma_V & \sigma_U^2 & \rho\sigma_U\sigma_V \\ \rho\sigma_U\sigma_V & \sigma_V^2 & \rho\sigma_U\sigma_V & \sigma_V^2 \\ \sigma_U^2 & \rho\sigma_U\sigma_V & \sigma_U^2 + s_{U,m}^2 & \rho\sigma_U\sigma_V \\ \rho\sigma_U\sigma_V & \sigma_V^2 & \rho\sigma_U\sigma_V & \sigma_V^2 + s_{V,m}^2 \end{pmatrix}.$$

Then, the conditional distribution of $(U_m, V_m)^{\mathrm{T}}$ given $(\widehat{U}_m, \widehat{V}_m)^{\mathrm{T}}$ is bivariate normal, with mean

$$\begin{pmatrix} \widetilde{\mu}_{U,m} \\ \widetilde{\mu}_{V,m} \end{pmatrix} = \begin{pmatrix} \mu_U \\ \mu_V \end{pmatrix} + AB_m^{-1}\begin{pmatrix} \widehat{U}_m - \mu_U \\ \widehat{V}_m - \mu_V \end{pmatrix}$$

and variance matrix

$$\begin{pmatrix} \Sigma_{11} & \Sigma_{12} \\ \Sigma_{12} & \Sigma_{22} \end{pmatrix} = A - AB_m^{-1}A,$$

where

$$A = \begin{pmatrix} \sigma_U^2 & \rho\sigma_U\sigma_V \\ \rho\sigma_U\sigma_V & \sigma_V^2 \end{pmatrix}, \quad B_m = \begin{pmatrix} \sigma_U^2 + s_{U,m}^2 & \rho\sigma_U\sigma_V \\ \rho\sigma_U\sigma_V & \sigma_V^2 + s_{V,m}^2 \end{pmatrix}.$$

Based on the proceeding derivations, for each step of the EM algorithm, we updated $(\mu_U, \mu_V, \sigma_U^2, \sigma_V^2, \rho)$ by

$$\begin{aligned}
\widehat{\mu}_U^{(j)} =& M^{-1}\sum_{m=1}^{M}\widetilde{\mu}_{U,m}, \qquad \widehat{\mu}_V^{(j)} = M^{-1}\sum_{m=1}^{M}\widetilde{\mu}_{V,m} \\
\widehat{\sigma}_U^{2,(j)} =& M^{-1}\sum_{m=1}^{M}(\widetilde{\mu}_{U,m}^2 + \Sigma_{11} - 2 * \widehat{\mu}_U^{(j)}\widetilde{\mu}_{U,m} + (\widehat{\mu}_U^{(j)})^2), \\
\widehat{\sigma}_V^{2,(j)} =& M^{-1}\sum_{m=1}^{M}(\widetilde{\mu}_{V,m}^2 + \Sigma_{22} - 2 * \widehat{\mu}_V^{(j)}\widetilde{\mu}_{V,m} + (\widehat{\mu}_V^{(j)})^2), \\
\widehat{\rho}^{(j)} =& \frac{M^{-1}\sum_{m=1}^{M}\widetilde{\mu}_{U,m}\widetilde{\mu}_{V,m} + \Sigma_{12} - \widehat{\mu}_U^{(j)}\widetilde{\mu}_{V,m} - \widehat{\mu}_V^{(j)}\widetilde{\mu}_{U,m} + \widehat{\mu}_U^{(j)}\widehat{\mu}_V^{(j)}}{\widehat{\sigma}_U^{(j)}\widehat{\sigma}_V^{(j)}},
\end{aligned}$$

where $\widetilde{\mu}_{U,m}, \widetilde{\mu}_{V,m}, \Sigma_{11}, \Sigma_{12}$, and $\Sigma_{22}$ are evaluated at the parameter value at the last step $(\widehat{\mu}_U^{(j-1)}, \widehat{\mu}_V^{(j-1)}, \widehat{\sigma}_U^{2,(j-1)}, \widehat{\sigma}_V^{2,(j-1)}, \widehat{\rho}^{(j-1)})$. We iterative between the E-step and M-step till the algorithm converges.

*Variance formulas for the counterfactual placebo HIV incidence and prevention efficacy*

Let $\widehat{\Sigma}$ be the estimated covariance matrix of $(\widehat{\mu}_U, \widehat{\mu}_V, \widehat{\sigma}_U^2, \widehat{\sigma}_V^2, \widehat{\rho})$ based on the maximum likelihood approach. By the delta method, the variance of $\widehat{U}_{0,k}$ can be estimated by

$$\widehat{var}(\widehat{U}_{0,k}) = c_1 \begin{pmatrix} \widehat{\Sigma} & 0 \\ 0 & \widehat{s}_{V,k}^2 \end{pmatrix} c_1^{\mathrm{T}},$$

where

$$c_1 = \left(1, -\frac{\widehat{\rho}\widehat{\sigma}_U\widehat{\sigma}_V}{\widehat{\sigma}_V^2 + \widehat{s}_{V,k}^2}, \frac{\widehat{\rho}\widehat{\sigma}_V}{2\widehat{\sigma}_U(\widehat{\sigma}_V^2 + \widehat{s}_{V,k}^2)}(\widehat{V}_k - \widehat{\mu}_V), \frac{\widehat{\rho}\widehat{\sigma}_U(\widehat{s}_{V,k}^2 - \widehat{\sigma}_V^2)}{2\widehat{\sigma}_V(\widehat{\sigma}_V^2 + \widehat{s}_{V,k}^2)^2}(\widehat{V}_k - \widehat{\mu}_V), \frac{\widehat{\rho}\widehat{\sigma}_U\widehat{\sigma}_V}{\widehat{\sigma}_V^2 + \widehat{s}_{V,k}^2}\right),$$

such that the variance of $\widehat{\lambda}_{0,k}^Y$ can be estimated by

$$\widehat{var}(\widehat{\lambda}_{0,k}^Y) = \{\widehat{\lambda}_{0,k}^Y(1 - \widehat{\lambda}_{0,k}^Y)\}^2\widehat{var}(\widehat{U}_{0,k})$$

by applying the delta method again. Then, the variance of $\widehat{PE}_k$ can be estimated by

$$\left(-\frac{1}{\widehat{\lambda}^Y_{0,k}}\right)^2 \widehat{var}(\widehat{\lambda}^Y_k) + \left(\frac{\widehat{\lambda}^Y_k}{(\widehat{\lambda}^Y_{0,k})^2}\right)^2 \widehat{var}(\widehat{\lambda}^Y_{0,k}) = \frac{1}{(\widehat{\lambda}^Y_{0,k})^2 n_{event,k}} + \frac{(1-\widehat{\lambda}^Y_{0,k})^2 \widehat{var}(\widehat{U}_{0,k})}{(\widehat{\lambda}^Y_{0,k})^2},$$

where $n_{event,k}$ is the total number of events observed in the arm $k$ in the trial.

*Supplemental Tables and Figures*

[Table 1 about here.]

[Figure 1 about here.]

[Figure 2 about here.]

[Figure 3 about here.]

[Table 2 about here.]

[Table 3 about here.]

[Table 4 about here.]

[Table 5 about here.]

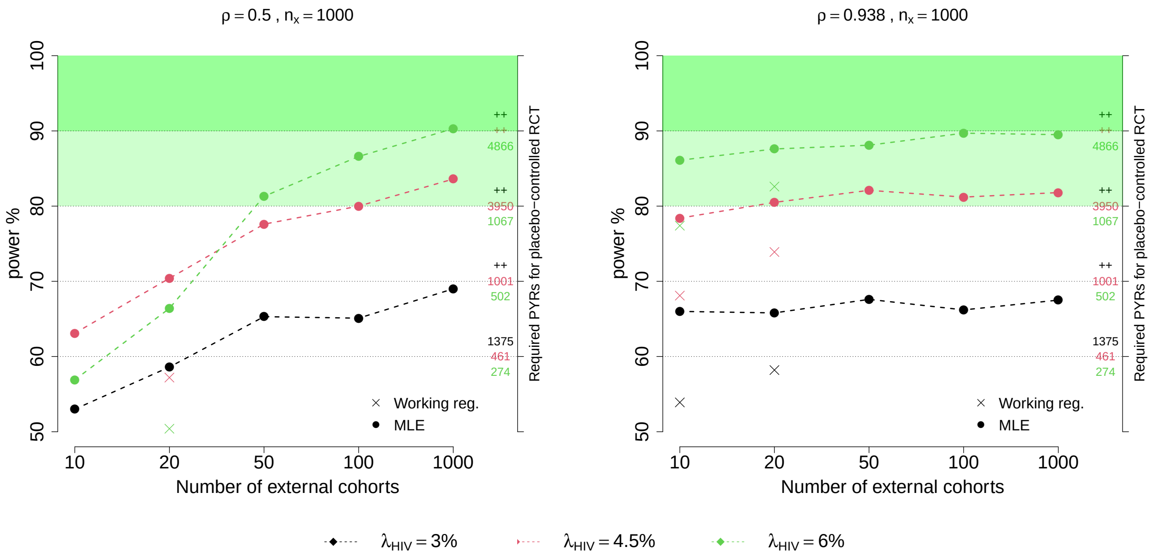


**Figure 1**: Power for testing $H_0: \ PE = 30\%$ vs. $H_a: \ PE = 60\%$ using the counterfactual approach as a function of $M$, the number of external cohorts. Given a fixed active arm size of 1000 person-years, the power based on estimating counterfactual placebo HIV incidence using MLE estimation, with a moderately correlated exposure marker ($\rho = 0.5$; left) or a highly correlated marker ($\rho = 0.938$; right). The size of a placebo arm, in person-years, required to obtain the corresponding power is shown on the right-hand y-axis. "++" indicates the required placebo arm sizes to reach the corresponding power is beyond 1,000,000 person-years. The power calculated from the working regression estimation approach is also shown for low $M$.

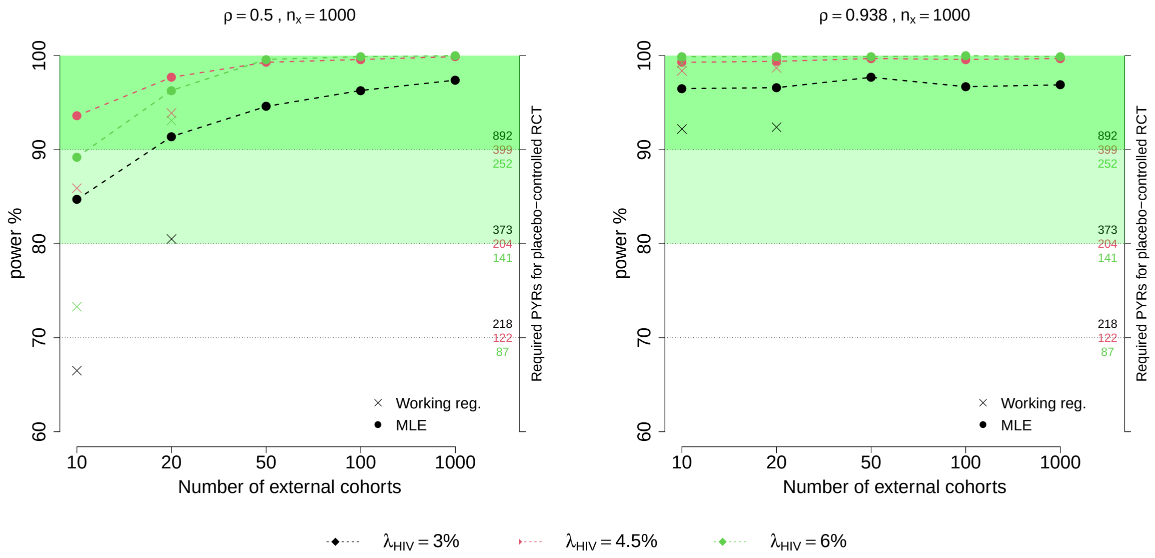


**Figure 2**: Power for testing $H_0: \ PE = 30\%$ vs. $H_a: \ PE = 75\%$ using the counterfactual approach as a function of $M$, the number of external cohorts. Given a fixed active arm size of 1000 person-years, the power based on estimating counterfactual placebo HIV incidence using MLE estimation, with a moderately correlated exposure marker ($\rho = 0.5$; left) or a highly correlated marker ($\rho = 0.938$; right). The size of a placebo arm, in person-years, required to obtain the corresponding power is shown on the right-hand y-axis. The power calculated from the working regression estimation approach is also shown for low $M$.

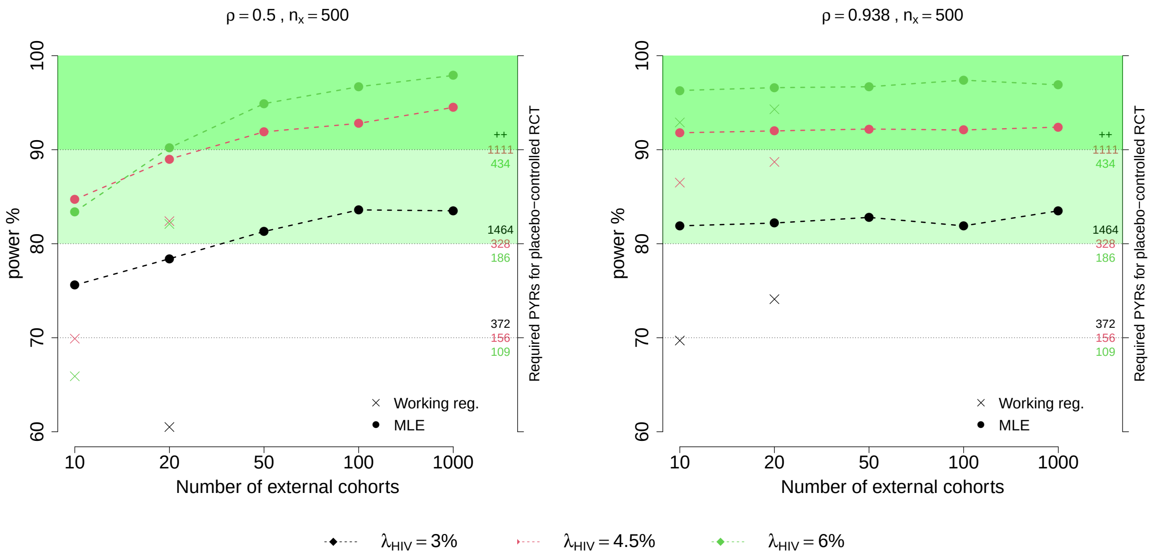


**Figure 3**: Power for testing $H_0:\ PE = 30\%$ vs. $H_a:\ PE = 75\%$ using the counterfactual approach as a function of $M$, the number of external cohorts. Given a fixed active arm size of 500 person-years, the power based on estimating counterfactual placebo HIV incidence using MLE estimation, with a moderately correlated exposure marker ($\rho = 0.5$; left) or a highly correlated marker ($\rho = 0.938$; right). The size of a placebo arm, in person-years, required to obtain the corresponding power is shown on the right-hand y-axis. "++" indicates the required placebo arm sizes to reach the corresponding power is beyond 1,000,000 person-years. The power calculated from the working regression estimation approach is also shown for low $M$

Table 1: Estimated HIV and RGC incidence rates for studies used as basis for simulations and analyzed in Mullick and Murray (2019)

| Referenced Study | HIV inc./100 PYRs | Num. PYRs | RGC inc./100 PYRs | Num. PYRs |
|---|---|---|---|---|
| Morris et al. (2006) | 2.5 | 943.2 | 3.5 | 943.2 |
| Jin et al. (2010) | 0.9 | 5160* | 2.3 | 5160* |
| Molina et al. (2015) | 6.6 | 212.1 | 15.5 | 212.1 |
| Castillo et al. (2015) | 3.6 | 1000* | 10.1 | 1000* |
| Kelley et al. (2015) | 3.8 | 843.1 | 6.2 | 726.6 |
| McGowan et al. (2016) | 6.4 | 50* | 16.1 | 50* |
| McCormack et al. (2016) | 9.0 | 245 | 33.1 | 596 |
| Girometti et al. (2017) | 8.3 | 100* | 33.0 | 100* |

*: person-years are approximate and not explicitly reported in the referenced studies.

Table 2: Percent bias, empirical coverage, and average 95% confidence intervals (CIs) for estimates of prevention efficacy (PE) based on $M$ external cohorts used to estimate the association between HIV and an exposure biomarker with correlation $\rho$. A total of $n_x = 1000$ person-years follow-up accrue in the active arm of the trial. Counterfactual placebo HIV incidence and true PE vary. Performance is shown for working model and likelihood-based estimation approaches.

| | $M$ | PE estimate | | $\rho = 0.938$ | | | $\rho = 0.5$ | |
|---|---|---|---|---|---|---|---|---|
| HIV incidence | | | 3% | 4.5% | 6% | 3% | 4.5% | 6% |
| Exposure marker incidence | | | 7.2% | 12.3% | 17.6% | 4.8% | 13.5% | 26.2% |
| | | | | Working model approach | | | | |
| $PE = 60\%$ | 10 | Bias% | -0.05 | -0.20 | -0.20 | 3.47 | 1.70 | 3.18 |
| $n_x = 1000$ | | Cov.% | 94.6 | 94.8 | 95.3 | 93.5 | 94.7 | 92.4 |
| | | AvgCI% | (29.5,81.0) | (36.2,77.4) | (39.1,75.7) | (-4.3,84.4) | (21.0,79.8) | (-33.0,80.4) |
| | 20 | Bias% | -0.32 | -0.43 | -0.45 | 1.03 | -0.25 | 0.12 |
| | | Cov.% | 94.0 | 95.1 | 94.7 | 94.4 | 94.2 | 93.6 |
| | | AvgCI% | (31.7,80.9) | (37.7,77.4) | (40.4,75.1) | (20.4,82.3) | (31.6,78.0) | (27.3,77.7) |
| $PE = 75\%$ | 10 | Bias% | -0.02 | -0.10 | -0.10 | 1.73 | 0.85 | 1.59 |
| $n_x = 1000$ | | Cov.% | 93.9 | 94.2 | 94.0 | 93.5 | 94.2 | 92.7 |
| | | AvgCI% | (52.6,91.2) | (57.0,88.4) | (59.2,86.9) | (31.4,92.4) | (47.7,89.4) | (14.5,89.0) |
| | 20 | Bias% | -0.16 | -0.21 | -0.23 | 0.52 | -0.12 | 0.06 |
| | | Cov.% | 93.3 | 94.2 | 93.9 | 94.2 | 93.4 | 93.9 |
| | | AvgCI% | (53.1,90.8) | (58.1,88.5) | (59.8,86.5) | (46.6,91.5) | (54.4,88.7) | (52.2,87.7) |
| | | | | Likelihood-based approach | | | | |
| $PE = 60\%$ | 20 | Bias% | -0.57 | 0.131 | -0.81 | -0.67 | -1.32 | -2.43 |
| $n_x = 1000$ | | Cov.% | 93.2 | 93.8 | 93.8 | 91.8 | 93.8 | 93.5 |
| | | AvgCI% | (36.0,83.4) | (40.8,79.3) | (42.6,76.4) | (31.9,87.3) | (38.2,81.3) | (35.2,81.9) |
| | 50 | Bias% | 0.00 | 0.07 | -0.64 | -0.14 | -0.10 | -0.55 |
| | | Cov.% | 92.95 | 93.9 | 93.6 | 92.1 | 93.4 | 94.0 |
| | | AvgCI% | (36.6,83.4) | (40.9,79.1) | (43.0,76.3) | (35.1,84.7) | (40.1,79.8) | (40.7,78.6) |
| $PE = 75\%$ | 20 | Bias% | -0.26 | -0.05 | -0.42 | -0.22 | -0.60 | -1.17 |
| $n_x = 1000$ | | Cov.% | 91.7 | 93.0 | 93.7 | 90.3 | 93.2 | 92.7 |
| | | AvgCI% | (56.5,93.1) | (60.0,89.9) | (61.6,87.8) | (54.4,95.2) | (58.1,91.0) | (57.5,90.8) |
| | 50 | Bias% | 0.05 | 0.16 | -0.30 | -0.08 | -0.09 | -0.35 |
| | | Cov.% | 92.1 | 93.3 | 92.9 | 90.9 | 92.7 | 93.1 |
| | | AvgCI% | (56.9,93.1) | (60.3,89.9) | (61.8,87.7) | (56.2,93.8) | (59.7,90.2) | (60.5,88.9) |

Table 3: Percent bias, empirical coverage, and average confidence intervals (CIs) for estimated counterfactual placebo HIV incidence, based on $M$ external cohorts used to estimate the association between HIV and an exposure biomarker with correlation $\rho$. A total of $n_x$ person-years follow-up accrue in the active arm of the trial. Counterfactual placebo HIV incidence varies. Performance is shown for working model and likelihood-based approaches. The conditional correlation $\rho_m$ are distributed uniformly in $(0.4, 0.5)$, while the estimation procedure assumes conditional independence.

| | | | | $\rho = 0.938$ | | | $\rho = 0.5$ | |
|---|---|---|---|---|---|---|---|---|
| HIV incidence | | | 3% | 4.5% | 6% | 3% | 4.5% | 6% |
| Exposure marker incidence | | | 7.2% | 12.3% | 17.6% | 4.8% | 13.5% | 26.2% |
| | | | | | Working model approach | | | |
| $M = 10$ | $n_x = 2000$ | Cov.% | 96.4 | 96.4 | 97.5 | 96.0 | 96.3 | 96.2 |
| | | Bias% | 0.04 | -0.05 | -0.00 | 3.37 | 1.54 | 4.22 |
| | | AvgCI% | (2.38,3.77) | (3.70,5.50) | (4.89,7.39) | (1.59,6.20) | (2.78,7.54) | (3.17,12.48) |
| | $n_x = 4000$ | Cov.% | 95.5 | 96.8 | 96.8 | 95.9 | 94.9 | 94.7 |
| | | Bias% | 0.15 | 0.19 | 0.35 | 2.88 | 2.21 | 6.22 |
| | | AvgCI% | (2.44,3.70) | (3.75,5.39) | (4.96,7.33) | (1.60,6.15) | (2.79,7.60) | (3.18,12.94) |
| $M = 20$ | $n_x = 2000$ | Cov.% | 93.6 | 95.8 | 97.5 | 95.2 | 96.5 | 95.1 |
| | | Bias% | 0.29 | 0.18 | 0.16 | 1.54 | 1.28 | 2.92 |
| | | AvgCI% | (2.53,3.57) | (3.91,5.21) | (5.20,6.92) | (1.98,4.68) | (3.30,6.28) | (3.97,9.56) |
| | $n_x = 4000$ | Cov.% | 95.4 | 96.2 | 97.0 | 95.1 | 95.3 | 95.6 |
| | | Bias% | 0.20 | 0.06 | 0.13 | 2.37 | 1.11 | 1.93 |
| | | AvgCI% | (2.60,3.47) | (3.97,5.10) | (5.27,6.83 | (2.01,4.68) | (3.32,6.23) | (3.95,9.41) |
| | | | | | Likelihood-based approach | | | |
| $M = 20$ | $n_x = 2000$ | Cov.% | 95.5 | 96.0 | 96.3 | 96.1 | 95.8 | 96.0 |
| | | Bias% | -2.03 | -1.528 | -0.88 | -1.57 | -1.15 | -0.51 |
| | | AvgCI% | (2.57,3.36) | (3.98,4.93) | (5.41,6.53) | (2.52,3.45) | (3.98,4.97) | (5.17,6.89) |
| | $n_x = 4000$ | Cov.% | 95.1 | 95.1 | 96.4 | 95.6 | 96.0 | 96.6 |
| | | Bias% | -1.89 | -1.39 | -0.83 | -1.83 | -1.18 | -0.49 |
| | | AvgCI% | (2.66,3.25) | (4.10,4.80) | (5.54,6.39) | (2.54,3.41) | (4.00,4.94) | (5.18,6.87) |
| $M = 50$ | $n_x = 2000$ | Cov.% | 94.8 | 95.0 | 94.8 | 95.5 | 94.1 | 95.2 |
| | | Bias% | -1.82 | -1.26 | -0.91 | -1.66 | -1.24 | -0.72 |
| | | AvgCI% | (2.60,3.34) | (4.03,4.90) | (5.46,6.47) | (2.64,3.30) | (4.11,4.80) | (5.44,6.52) |
| | $n_x = 4000$ | Cov.% | 93.9 | 93.7 | 94.7 | 94.4 | 93.9 | 95.2 |
| | | Bias% | -2.03 | -1.47 | -0.84 | -1.83 | -1.19 | -0.55 |
| | | AvgCI% | (2.68,3.22) | (4.13,4.76) | (5.59,6.33) | (2.67,3.25) | (4.15,4.77) | (5.47,6.51) |

Table 4: Percent bias, empirical coverage, and average 95% confidence intervals (CIs) for estimates of prevention efficacy (PE) based on $M$ external cohorts used to estimate the association between HIV and an exposure biomarker with correlation $\rho$. A total of $n_x = 2000$ person-years follow-up accrue in the active arm of the trial. Counterfactual placebo HIV incidence and true PE vary. Performance is shown for working model and likelihood-based approaches. The conditional correlation $\rho_m$ are distributed uniformly in $(0.4, 0.5)$, while the estimation procedure assumes conditional independence.

| | $M$ | PE estimate | | $\rho = 0.938$ | | | $\rho = 0.5$ | |
|---|---|---|---|---|---|---|---|---|
| HIV incidence | | | 3% | 4.5% | 6% | 3% | 4.5% | 6% |
| Exposure marker incidence | | | 7.2% | 12.3% | 17.6% | 4.8% | 13.5% | 26.2% |
| | | | | Working model approach | | | | |
| $PE = 30\%$ | 10 | Bias% | 1.16 | 0.44 | 0.25 | 7.60 | 3.53 | 9.45 |
| $n_x = 2000$ | | Cov.% | 93.9 | 94.1 | 93.8 | 92.8 | 93.0 | 90.8 |
| | | AvgCI% | (-0.8,52.0) | (6.0,48.6) | (8.2,47.7) | (-66.4,62.2) | (-21.8,56.2) | (-60.2,60.4) |
| | 20 | Bias% | 0.67 | 0.41 | 0.37 | 3.66 | 3.02 | 6.66 |
| | | Cov.% | 95.2 | 95.9 | 94.9 | 95.0 | 94.4 | 92.5 |
| | | AvgCI% | (2.7,51.4) | (8.5,47.7) | (10.6,45.5) | (-20.9,57.2) | (-3.3,52.4) | (-14.2,54.8) |
| $PE = 60\%$ | 10 | Bias% | 0.03 | -0.03 | 0.00 | 2.17 | 1.01 | 2.70 |
| $n_x = 2000$ | | Cov.% | 95.5 | 95.4 | 94.8 | 93.0 | 93.4 | 92.4 |
| | | AvgCI% | (38.8,75.8) | (43.4,73.2) | (45.0,72.1) | (2.2,80.1) | (28.0,76.5) | (6.8,78.3) |
| | 20 | Bias% | 0.19 | 0.12 | 0.11 | 1.01 | 0.84 | 1.89 |
| | | Cov.% | 95.5 | 94.9 | 95.0 | 95.3 | 93.4 | 92.8 |
| | | AvgCI% | (40.6,75.5) | (44.7,72.9) | (46.2,71.1) | (27.9,77.8) | (38.4,74.8) | (32.8,75.3) |
| $PE = 75\%$ | 10 | Bias% | 0.02 | -0.02 | -0.01 | 1.10 | 0.46 | 1.28 |
| $n_x = 2000$ | | Cov.% | 94.3 | 94.6 | 94.2 | 93.0 | 94.6 | 93.5 |
| | | AvgCI% | (59.1,87.0) | (62.2,84.9) | (64.0,84.2) | (36.9,88.9) | (52.9,86.5) | (40.2,87.3) |
| | 20 | Bias% | 0.12 | 0.07 | 0.05 | 0.57 | 0.43 | 0.94 |
| | | Cov.% | 94.1 | 93.3 | 95.2 | 94.8 | 94.6 | 93.4 |
| | | AvgCI% | (60.4,87.0) | (63.0,84.7) | (64.6,83.6) | (53.0,88.0) | (59.4,85.6) | (56.7,85.6) |
| | | | | Likelihood-based approach | | | | |
| $PE = 30\%$ | 20 | Bias% | -0.78 | -0.16 | -0.75 | -4.32 | -2.20 | -5.01 |
| $n_x = 2000$ | | Cov.% | 94.4 | 94.0 | 94.5 | 92.4 | 93.1 | 92.5 |
| | | AvgCI% | (6.3,53.2) | (10.9,49.0) | (13.0,46.6) | (-5.9,63.3) | (3.0,55.7) | (-3.1,60.1) |
| | 50 | Bias% | 0.40 | 0.15 | 0.09 | 0.29 | -0.63 | -2.31 |
| | | Cov.% | 94.3 | 94.6 | 94.9 | 93.6 | 94.3 | 94.0 |
| | | AvgCI% | (7.3,52.9) | (11.5,48.6) | (13.9,46.1) | (2.8,57.4) | (8.4,51.2) | (6.3,52.4) |
| $PE = 60\%$ | 20 | Bias% | -0.39 | -0.06 | -0.14 | -1.41 | -0.66 | -1.35 |
| $n_x = 2000$ | | Cov.% | 94.3 | 93.6 | 94.9 | 92.4 | 93.0 | 92.8 |
| | | AvgCI% | (42.8,76.8) | (46.1,73.8) | (47.8,72.1) | (36.5,81.8) | (42.1,77.1) | (39.3,79.1) |
| | 50 | Bias% | 0.15 | -0.09 | 0.05 | 0.07 | -0.34 | -0.59 |
| | | Cov.% | 94.5 | 94.6 | 94.5 | 93.7 | 94.2 | 94.0 |
| | | AvgCI% | (43.4,76.8) | (46.3,73.6) | (48.2,71.9) | (41.2,78.9) | (44.8,74.8) | (44.4,74.9) |
| $PE = 75\%$ | 20 | Bias% | -0.28 | 0.00 | -0.11 | -0.79 | -0.29 | -0.72 |
| $n_x = 2000$ | | Cov.% | 93.3 | 93.9 | 94.4 | 91.9 | 93.1 | 92.8 |
| | | AvgCI% | (61.7,87.9) | (64.3,85.7) | (65.5,84.3) | (58.2,90.6) | (62.1,87.4) | (60.8,88.2) |
| | 50 | Bias% | 0.15 | -0.16 | 0.00 | 0.08 | -0.29 | -0.31 |
| | | Cov.% | 93.5 | 94.4 | 94.2 | 93.9 | 93.9 | 94.0 |
| | | AvgCI% | (62.2,88.0) | (64.3,85.5) | (65.8,84.2) | (61.0,89.1) | (63.4,86.1) | (63.7,85.8) |

Table 5: A comparison of cohort-level vs. site-level inference. Percent bias, empirical coverage, and average confidence intervals (CIs) for estimated counterfactual placebo HIV incidence and PE, based on $M = 20$ external cohorts used to estimate the association between HIV and an exposure biomarker with correlation $\rho$. A total of $n_x = 2000$ person-years follow-up accrue in the active arm of the trial. Counterfactual placebo HIV incidence varies. Performance is shown for working model and likelihood-based approaches. Each of $M = 20$ cohorts have $L = 5$ sites and the inference is compared based on analysis of $M = 20$ cohorts vs. $M \times L = 100$ sites.

| | | | $\rho = 0.938$ | | | $\rho = 0.5$ | | |
|---|---|---|---|---|---|---|---|---|
| HIV incidence | | | 3% | 4.5% | 6% | 3% | 4.5% | 6% |
| Exposure marker incidence | | | 7.2% | 12.3% | 17.6% | 4.8% | 13.5% | 26.2% |
| Est. counterfactual placebo HIV incidence | | | | | | | | |
| | | | Working model approach | | | | | |
| $M = 20$ | Cohort-level | Cov.% | 94.6 | 96.1 | 96.3 | 95.4 | 93.4 | 94.6 |
| | | Bias% | -0.61 | -0.71 | -0.68 | 0.50 | 0.38 | 1.96 |
| | | AvgCI% | (2.49,3.57) | (3.85,5.20) | (5.12,6.94) | (1.95,4.65) | (3.27,6.22) | (3.93,9.49) |
| $M \times L = 100$ | Site-level | Cov.% | 93.8 | 93.5 | 94.2 | 93.0 | 92.5 | 94.3 |
| | | Bias% | -0.28 | -2.1 | -3.31 | 0.83 | -1.53 | -2.81 |
| | | AvgCI% | (2.60,3.45) | (3.91,4.94) | (5.18,6.49) | (2.46,3.69) | (3.81,5.15) | (4.77,7.10) |
| | | | Likelihood-based approach | | | | | |
| $M = 20$ | Cohort-level | Cov.% | 95.5 | 95.5 | 95.1 | 94.5 | 94.3 | 94.5 |
| | | Bias% | 0.43 | 0.18 | 0.01 | 1.97 | 1.13 | 2.36 |
| | | AvgCI% | (2.57,3.53) | (3.96,5.13) | (5.30,6.79) | (2.05,4.57) | (3.38,6.12) | (4.08,9.21) |
| $M \times L = 100$ | Site-level | Cov.% | 94.9 | 95.5 | 94.5 | 95.1 | 94.8 | 94.9 |
| | | Bias% | 0.73 | 0.28 | 0.22 | 1.50 | 0.78 | 0.71 |
| | | AvgCI% | (2.66,3.44) | (4.08,4.99) | (5.50,6.57) | (2.52,3.67) | (3.96,5.20) | (5.05,7.22) |
| Est. PE, PE=60% | | | | | | | | |
| | | | Working model approach | | | | | |
| M=20 | Cohort-level | Cov.% | 95.7 | 94.9 | 95.4 | 94.8 | 94.7 | 94.8 |
| | | Bias% | -0.41 | -0.47 | -0.46 | 0.33 | 0.25 | 1.28 |
| | | AvgCI% | (39.6,75.3) | (44.1,72.7) | (46.2,71.4) | (27.0,77.7) | (38.0,74.6) | (33.1,75.7) |
| $M \times L = 100$ | Site-level | Cov.% | 95.1 | 93.1 | 94.5 | 93.6 | 95.2 | 94.7 |
| | | Bias% | -0.77 | -1.88 | -2.81 | 0.06 | -1.65 | -2.49 |
| | | AvgCI% | (40.4,75.0) | (43.5,71.9) | (45.0,69.9) | (39.3,75.8) | (42.9,72.3) | (42.5,71.1) |
| | | | Likelihood-based approach | | | | | |
| $M = 20$ | Cohort-level | Cov.% | 93.4 | 94.8 | 94.8 | 92.5 | 93.5 | 93.4 |
| | | Bias% | -0.32 | -0.01 | -0.36 | -1.54 | -0.91 | -1.35 |
| | | AvgCI% | (42.8,76.8) | (46.1,73.8) | (47.6,72.0) | (36.3,81.8) | (41.9,77.0) | (39.3,79.1) |
| $M \times L = 100$ | Site-level | Cov.% | 93.2 | 94.2 | 94.4 | 93.6 | 93.9 | 94.6 |
| | | Bias% | -0.06 | -0.01 | -0.12 | 0.32 | 0.18 | -0.14 |
| | | AvgCI% | (43.3,76.6) | (46.4,73.5) | (48.2,71.7) | (42.7,77.7) | (46.1,74.1) | (46.5,73.3) |
